# Racial and Ethnic Disparities in Brain Age Algorithm Performance: Investigating Bias Across Six Popular Methods

**DOI:** 10.1101/2025.09.18.25336117

**Authors:** Dorthea J Adkins, Jamie L. Hanson

## Abstract

Brain age algorithms, which estimate biological aging from neuroimaging data, are increasingly used as biomarkers for health and disease. However, most algorithms are trained on datasets with limited racial and ethnic diversity, raising concerns about potential algorithmic bias that could exacerbate health disparities. To probe this potential, we evaluated six popular brain age algorithms using data from the Health and Aging Brain Study–Health Disparities (HABS-HD), comprising 1,123 White American, 1,107 Hispanic American, and 678 African American participants, ages ≥50. Comparing correlations between brain age and chronological age across racial/ethnic groups, relations were consistently weaker for African American participants compared to White and Hispanic American participants across most algorithms (ranging from r=0.51-0.85 for African Americans vs. r=0.57-0.89 for other groups). We also examined error for brain age v. chronological age and found significant differences in median errors across racial/ethnic groups, though specific patterns varied by algorithm. Sensitivity models weighting for age, sex, and scan quality noted similar patterns, with all algorithms maintaining significant differences in correlation or median prediction error between groups. Our findings reveal systematic performance differences in brain age algorithms across racial and ethnic groups, with most algorithms consistently showing reduced algorithm accuracy for African American and/or Hispanic-American participants. These biases, which are likely introduced at multiple stages of algorithm development, could impact clinical utility and diagnostic accuracy. Results highlight the urgent need for more inclusive algorithm development and validation to ensure equitable healthcare applications of neuroimaging biomarkers.

## INTRODUCTION

While the causes of aging remain poorly understood, its impact is clear. A growing proportion of the global population shows age-associated functional decline and disease (Z. Li et al., 2021). To better understand pathways to longevity and age-related deteriorations, researchers increasingly focus on “biological age” and its divergence from chronological age. These approaches examine telomeres, epigenetics, and other molecular and cellular markers to provide more precise measures of morbidity and mortality risk (Mather et al., 2011; Pal & Tyler, 2016).

Novel biological age metrics have emerged by applying machine learning algorithms to neuroimaging data to calculate “brain age.” By leveraging large datasets containing age and neuroimaging scans, researchers can estimate brain age in new participants using only neuroimaging data. These estimates, typically derived from structural MRI scans, often diverge from chronological age, suggesting alterations in biological aging. This divergence is significant because aging affects individuals non-uniformly, both within and between people. Given the brain’s central role in regulating behavior and neuroendocrine processes, brain age metrics may serve as particularly predictive indicators of age-related mortality and morbidity. This has led to a rapid rise in research groups using brain age, with this metric being employed as a potential biomarker for neurodegeneration, cognitive decline, and multiple issues of importance to public health (e.g., schizophrenia, major depression, bipolar disorder (Baecker et al., 2021).

While focusing on brain age could enable more comprehensive models of biological aging and biomarkers for predicting important clinical outcomes (Cole et al., 2019), there is variability in how these algorithms are developed and implemented. As reviewed by (Dörfel et al., 2023), there are variations in the data analytic techniques (e.g., Convolutional Neural Network; Ridge Regression, Gaussian Process Regression), the brain features used (e.g., derived regions of interest; MRI voxels), and the data used for training. This latter element may be of particular concern as many brain age algorithms are trained on well-established MRI datasets like the UK Biobank, the Cambridge Centre for Ageing Neuroscience dataset, Alzheimer’s Neuroimaging Initiative, the Open Access Series of Imaging Studies, and other projects (Piçarra & Glocker, 2023). Many of these publicly available datasets tend to be demographically skewed, especially in terms of ethnic and racial diversity, with Non-hispanic White subjects often composing the near totality of participants.

The composite of an algorithm’s training dataset may have major implications for its ability to be widely applied, especially to ethnic and racial minority groups and other populations underrepresented in research. For example, when detecting skin cancer, cutting edge machine learning approaches were initially found to perform on par or superior to dermatologists (Esteva et al., 2017); however, further investigations showed that these algorithms perform quite poorly on dark skin tones and uncommon diseases (Daneshjou et al., 2022; Kleinberg et al., 2022). Notably, there have been suggestions of potential racial or ethnic biases in the quantification of biological age, such as epigenetic age (Philibert et al., 2020). Put another way, if developed without diverse representation, biological age calculators could potentially miscalculate age for individuals in marginalized groups, which may have downstream negative consequences for their clinical utility and increase health disparities (Chen et al., 2023). Related to brain age and how these algorithms are typically developed using data primarily from Non-hispanic White subjects, it will be critical to examine if different commonly used brain age algorithms generate comparable predictions for white and non-white participants. This is particularly important given that brain aging is being investigated in diverse cohorts and divergent patterns of brain aging are being reported between different racial and ethnic groups (Dempsey et al., 2023; Turney et al., 2023). No study, to our knowledge, has examined if commonly-used brain age algorithms exhibit potential bias in racial and ethnic minority groups.

To fill these gaps, here we leveraged a publically accessible dataset of White, Black, and Hispanic participants and compared brain age prediction between groups. Processing these datasets through six commonly-used brain age algorithms (4 algorithms using Freesurfer-derived outputs; 2 algorithms using less processed T1-weighted MRI scans), we first calculated the correlations between brain age and chronological age for each participant and algorithm, and tested for differences in these correlations between racial and ethnic groups. We then examined group differences in algorithmic error using non-parametric ANOVA methods. Given past work finding systematic biases for racial or ethnic minority groups in medical algorithm prediction, we predicted that the accuracy for Non-hispanic White subjects would be superior to Black and Hispanic participants for a number of commonly-used brain age algorithms. To address potential confounding variables, we also implemented a propensity score weighting approach as a secondary analysis. This allowed for balancing key demographic characteristics across groups, thereby isolating the effects of race/ethnicity on algorithm performance from other potentially confounding factors.

## METHODS

### Participants

The Health and Aging Brain Study–Health Disparities (HABS-HD) is an ongoing study that aims to discern the factors underlying racial/ethnic differences in rates of Alzheimer’s Disease and develop targeted medical interventions for this disorder (Clark et al., 2024; Hall et al., 2022; O’Bryant et al., 2021). First established in 2017 as the Health and Aging Brain among Latino Elders study, the HABS-HD study was approved by the University of North Texas Health Science Center Institutional Review Board. While the project initially aimed to elucidate the causes of differences in neurocognitive outcomes between Mexican Americans and non Hispanic Whites only, the study now also recruits African-Americans and currently aims to recruit 1,000 participants for each racial group (Hall et al., 2022; O’Bryant et al., 2021). HABS-HD participants are currently required to be adults aged 50 or older, to be fluent in Spanish or English, and identify as Black/African American, Hispanic, or Non-Hispanic White (O’Bryant et al., 2021). Included participants are also required to be eligible to undergo brain scanning. Participants are excluded from HABS-HD if they have a severe medical or mental illness (other than depression) with the potential for effects on cognition and/or traumatic brain injury with a loss of consciousness within the past year. All study participants give written informed consent.

For the current study, to minimize overlap between the groups, participants who endorsed both “White” and “Black” (N = 2) were excluded. Hispanic participants who endorsed “Black” (N = 8) and/or non-Mexican Hispanic ancestry (N = 12), as well as those who did not endorse “White” (N = 16) were also excluded. The sample was further filtered to exclude participants with Euler numbers greater than 100 (N = 24) as well as participants whose MRI scans had CAT12 grades less than .8 (N = 4). Euler numbers and CAT12 grades are metrics of MRI scan quality. The final sample consisted of a total of 2907 participants, of which 1123 were White American, 1106 were Hispanic American, and 678 were African American. Table 1 details the study’s demographics.

**Table 1.** Tabulated here are the sample sizes, the mean and standard deviation of age and Euler Number, and the numbers and percentages of each sex, for each racial/ethnic group.

| Age, Sex, and Euler Number by Group |  |  |  |
| --- | --- | --- | --- |
| Characteristic | Black<br>N = 678<sub>1</sub> | Hispanic<br>N = 1,106<sub>1</sub> | White<br>N = 1,123<sub>1</sub> |
| Age | 62.83 (7.84) | 63.22 (8.06) | 68.53 (8.69) |
| Sex |  |  |  |
| F | 439 / 678 (65%) | 740 / 1,106 (67%) | 642 / 1,123 (57%) |
| M | 239 / 678 (35%) | 366 / 1,106 (33%) | 481 / 1,123 (43%) |
| Euler Number | 21.79 (15.19) | 17.87 (11.30) | 21.67 (14.02) |

<sub>1</sub>Mean (SD); n / N (%)

### Brain Age Algorithms

We deployed six brain age algorithms on this dataset: (Cole et al., 2018) (referred to as “brainageR”), (Kaufmann et al., 2019) (referred to as “XGBoost”), (Bashyam et al., 2020) (referred to as “DeepBrainNet”), (Han et al., 2021) (referred to as “ENIGMA”), (Leonardsen et al., 2022) (referred to as “Pyment”); and (Yin et al., 2023) (referred to as “USC”). We selected these algorithms based on popularity in recent brain age publications and their open access code. For DeepBrainNet and brainageR, T1-weighted MRI scans are preprocessed using SPM12 (brainageR) or multi-atlas label fusion and FMRIB’s FLIRT (DeepBrainNet) before input into each algorithm. In contrast, Pyment, XGBoost, ENIGMA, and USC require preprocessing using Freesurfer (Fischl, 2012), an open-source MRI processing software package completed using brainlife.io (Hayashi et al., 2024). We provide brief summaries of each algorithm below. For detailed descriptions of model structure, please see the original papers cited here.

#### *brainageR Algorithm* (Cole et al., 2018)

brainageR loaded T1-weighted MRI images into SPM12 where these anatomical images were bias-corrected, segmented and normalized with custom brain templates. After this, the resulting segmented and normalized images were loaded into R (Cole et al., 2018), where gray matter, white matter, and CSF were then vectorized. The rotation matrix of a previously calculated Principal Components Analysis was then applied to these vectors to predict age values with *kernlab* (using a Gaussian Process Regression with Radial Basis Function kernel and default hyperparameters). This algorithm (version 2.0, 24 Sep 2019) was trained on a sample (N=2001) of healthy adults aged 18-90. Relevant code is available at: https://github.com/james-cole/brainageR.

#### *DeepBrainNet Algorithm* (Bashyam et al., 2020)

DeepBrainNet is a 2D Convolutional Neural Network (CNN) built using the inception-resnetv2 framework and pre-trained on ImageNet (Bashyam et al., 2020). With this algorithm, raw, unprocessed, T1-weighted MR images are N4 bias corrected, skull-stripped, and affine registered with FMRIB’s FLIRT to a common atlas space. This algorithm was implemented through the ANTsRNet package, an implementation of Advanced Normalization Tools (ANTs) in the R programming language (Tustison et al., 2021). This algorithm was trained on a sample (N = 11,729) of healthy controls aged 3-95. Relevant code for this algorithm is located here: https://github.com/ANTsX/brainAgeR.

#### *XGBoost Algorithm* (Kaufmann et al., 2019)

XGBoost uses gradient tree boosting to predict brain age based on 1118 features extracted using Freesurfer (Kaufmann et al., 2019), such as thickness, area, and volume measurements (Glasser et al., 2016). This algorithm was trained on a large and diverse sample (N=35,474, ages 3-89) as well as trained separately for male and female brain age. We deployed this algorithm by first completing standard processing approaches in Freesurfer 7.1 (http://surfer.nmr.mgh.harvard.edu), which includes motion correction and intensity normalization of T1-weighted images; removal of non-brain tissue; automated Talairach transformation; segmentation of white matter and gray matter volumetric structures; and derivation of cortical thickness. Relevant code for the XGBoost algorithm is available at: https://github.com/tobias-kaufmann/brainage

#### *ENIGMA Algorithm* (Han et al., 2021)

The ENIGMA algorithm used ridge regression based on Freesurfer features (as noted above) and was developed based on data from N = 2,188 (aged 18-75) participants (Han et al., 2021). Notably, these data were split within each of 19 scanning sites into training and test sets with equivalent chronological age distributions. Freesurfer-derived structural MRI measures from both the left and right hemispheres were combined to create 77 brain features, including subcortical volumes, cortical thickness, and surface area. Normative models were then estimated using ridge regression in a training sample of male and female controls, with brain age estimated separately for each sex. Relevant code for this algorithm is located here: https://photon-ai.com/enigma_brainage.

#### *Pyment Algorithm* (Leonardsen et al., 2022)

The Pyment algorithm implemented a Simple Fully Convolutional Network on T1-weighted structural magnetic resonance images. These images were partially preprocessed in FreeSurfer (using this software’s auto-recon steps) before being reoriented to the standard FSL and registered linearly using FLIRT. The training dataset was one of the largest and most diverse datasets assembled (N= 53,542, ages 3-95), stratified by age and study. Notably, data from the older participants (>40 years) in the training set was primarily derived from the UK Biobank. Additional technical details are available in the original report (Ref. (Leonardsen et al., 2022)). Relevant code for this algorithm is located here: https://github.com/estenhl/pyment-public

#### *USC Algorithm* (Yin et al., 2023)

The USC algorithm leverages an interpretable three-dimensional convolutional neural network (CNN) to estimate brain age based on T1-weighted MRI images. This algorithm was trained and tested on fMRI data from N = 4,681 (22-95 y) cognitively normal participants from four independent samples, with most data derived from the UK Biobank. All MRIs were reconstructed and segmented in FreeSurfer using the recon-all function, and registered to the MNI305 atlas. MRIs were then input into a deep learning regression model that utilizes three convolutional blocks followed by two dense layers to generate brain ages. Relevant code for this algorithm is located here: https://github.com/irimia-laboratory/USC_BA_estimator

### MRI Image Quality Assessment

To assess structural image quality, we calculated the Euler number for each participant using FreeSurfer’s cortical surface reconstruction output. The Euler number is a quantitative measure of topological complexity of the reconstructed cortical surface, calculated separately for each hemisphere and averaged to produce a single value per participant. In the present study, we were motivated to account for this metric because: (1) Euler Number and Freesurfer outputs such as cortical thickness are correlated with MRI quality (Gilmore et al., 2021; Rosen et al., 2018) and (2) brain age is correlated with MRI quality even after removing poor-quality structural images (Bacas et al., 2023; Hanson et al., 2024).

### Statistical Analyses

To understand potential biases in the calculation of brain age for different racial and ethnic groups, we deployed tests of differences between independent correlations to compare chronological age and brain age correlations for each algorithm and for each racial/ethnic group. After dividing the sample by race/ethnicity, we first computed bivariate CA-BA correlations for each brain age algorithm and for each group (6 algorithms x 3 ethnic groups = 18 total correlations). Separately for each algorithm, Fisher’s z and Zou’s tests (α = 0.05) were then utilized to compare the chronological age and brain age correlations between: (a) White- and African-American; (b) White- and Hispanic-American; and (c) Hispanic- and African-American.

Next, we probed differences in error for each algorithm between racial/ethnic groups. Of note, this was calculated as the raw prediction errors (predicted - actual) without an absolute or squared value transformation. Given concerns about algorithmic bias in medical applications and our hypothesis that brain age algorithms may systematically over- or under-predict age for different demographic groups, it was critical for our comparison to preserve positive or negative errors in predicted values. Other common performance metrics (e.g., Mean Absolute Error; Root Mean Squared Error) would obscure over-prediction and under-prediction as equivalent failures. For each algorithm, Kruskal-Wallis rank sum tests (α = 0.05) were deployed to compare the median errors of the racial/ethnic groups. This was used because the characteristics of the dataset violated the assumptions of a parametric ANOVA. If a Kruskal-Wallis rank sum test was significant, post-hoc Dunn’s tests were used to identify which groups differed significantly by median error.

As an additional sensitivity analysis and to limit potential demographic confounding, propensity scores were estimated with age, sex, and Euler number in relation to race/ethnicity. These propensity scores were then used to calculate inverse probability weights, allowing for the computation of the average treatment effect of race/ethnicity in subsequent statistical models while retaining all participants in the analysis. Using the propensity score weights, we first computed weighted correlations between brain and chronological age separately for each group and algorithm (ANTS, brainageR, ENIGMA, Kaufmann, Pyment, USC). These weighted correlations were then compared across groups for each algorithm, using the same analyses that were utilized in the full sample. We then conducted weighted Kruskal-Wallis tests for overall group differences in brain age prediction errors for each algorithm. Because weighted Dunn’s tests were not available in R, significant weighted Kruskal-Wallis tests were instead followed by weighted pairwise Wilcoxon tests.

## RESULTS

### Analyses in Full Sample

Correlations between brain age and chronological age for each racial and ethnic group are shown in Table 2 and Figure 1. These ranged from r=.58-.89 for White participants, r=.51-.85 for African American participants, and r=.57-.88 for Hispanic participants.

**Table 2.**
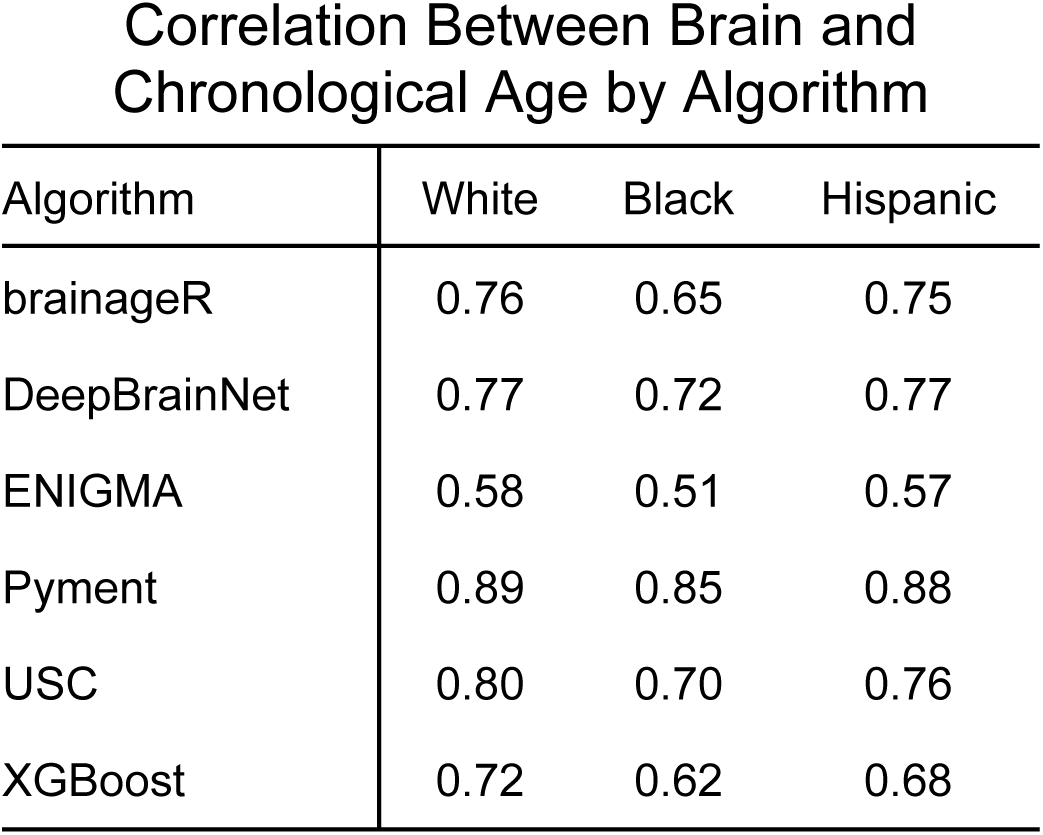
Bivariate correlations between brain age and chronological age were calculated separately for each algorithm and racial/ethnic group.

**Figure 1:**
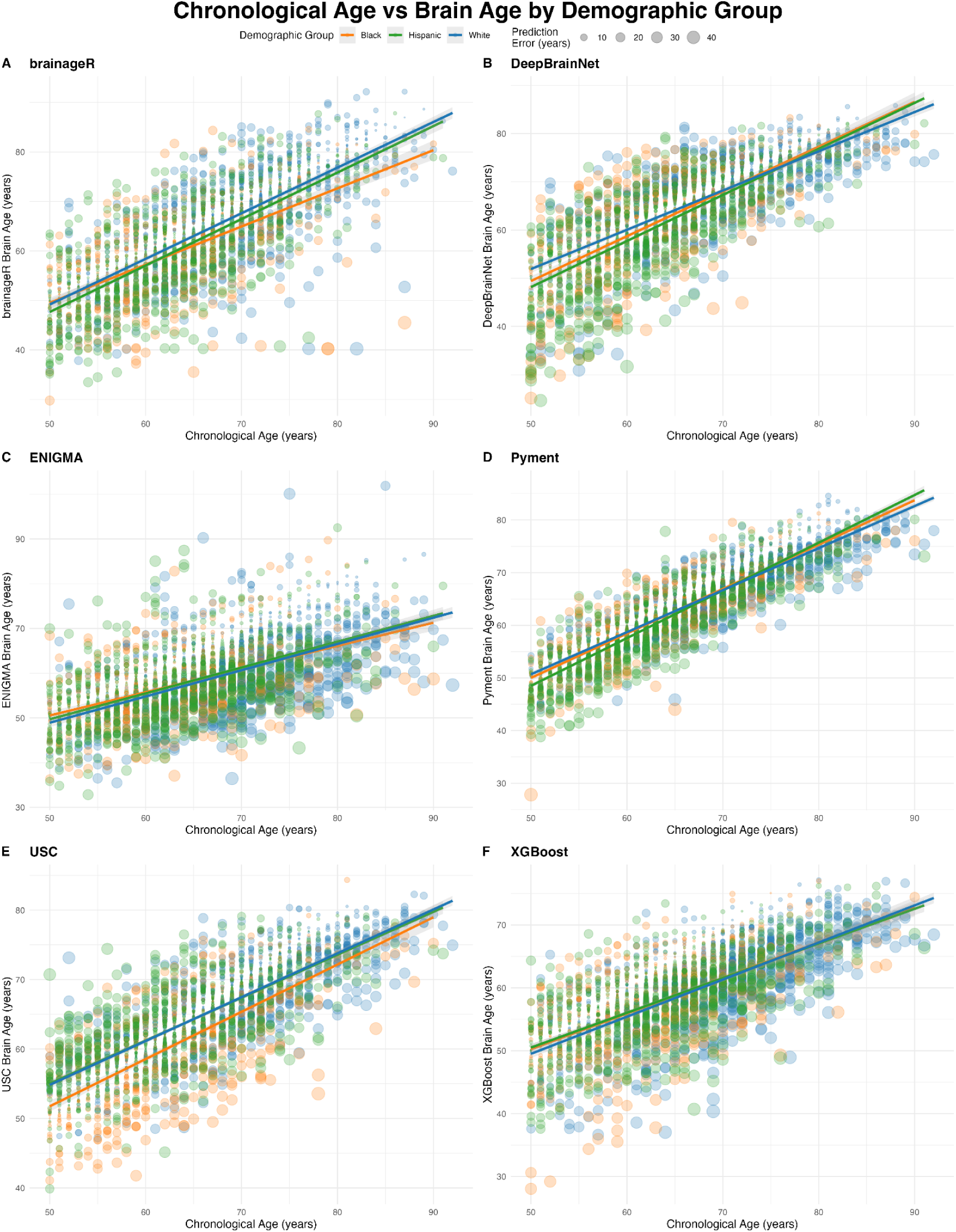
Scatterplots depicting the linear regression line for WA (Blue), AA (Orange) and MA (Green) for brainageR (A), DeepBrainNet (B), ENIGMA (C), Pyment (D), USC (E), and XGBoost (F). Points are sized by the magnitude of the prediction error.

Across algorithms, correlations between brain age and chronological age were often significantly different when comparing African American participants to White or Hispanic American participants. When comparing White and African American participants, five of the six algorithms showed significantly different correlations between brain age and chronological age. Specifically, differences were found for brainageR (p-uncorrected < 0.001, p-fdr < 0.001, z = 4.70, CI = 0.065-0.166), XGBoost (p-uncorrected < 0.001, p-fdr < 0.001, z = 3.81, CI = 0.049-0.158), Pyment (p-uncorrected = 0.002, p-fdr=0.008, z = 3.05, CI = 0.013-0.061), DeepBrainNet (p-uncorrected = 0.009, p-fdr=0.022, z = 2.63, CI = 0.014-0.101), and USC (p-uncorrected < 0.001, p-fdr < 0.001, z = 4.63, CI = 0.055-0.142).

Comparing African American and Hispanic participants, five of six algorithms showed differences (USC: p-uncorrected= 0.023, p-fdr=0.043, z = 2.28, CI = 0.007-0.099; XGBoost: p-uncorrected = 0.027, p-fdr= 0.043, z = 2.22, CI = 0.007-0.120; Pyment: p = 0.024, p-fdr=0.043, z = 2.26, CI = 0.004-0.053; brainageR: p-uncorrected < 0.001, p-fdr < 0.001, z = 4.16, CI = 0.053-0.155; DeepBrainNet: p = 0.025, p-fdr= 0.043, z = 2.25, CI = 0.006-0.094).

Lastly, examining White and Hispanic participants, only USC showed significantly different correlations between brain age and chronological age (p-uncorrected = 0.007, p-fdr= 0.021, z = 2.70, CI = 0.012-0.078). No significant differences between White Americans and Hispanic Americans were found between the correlation coefficients for the other algorithms (z=0.123-1.823). Notably, differences were not seen between any of the groups for the ENIGMA algorithm (z=0.123-1.86), but correlations between brain age and chronological age were also the lowest for this algorithm (r=.51-.58).

In addition to examining correlations between brain age and chronological age, we also probed the performance error by subtracting chronological age from brain age for each individual and each algorithm. Figure 2 shows the distributions of median errors by race and ethnicity for each algorithm, while Table 3 contains the median errors for all algorithms and groups. For all algorithms investigated, Kruskal-Wallis tests indicated that the median errors of the racial/ethnic groups significantly differed from one another (brainageR: H = 16.19, p < .001; DeepBrainNet: H = 13.5, p = 0.0012; ENIGMA: H = 106.9, p < 0.001; Pyment: H = 29.96, p < 0.001; USC: H = 103.02, p = < 0.001; XGBoost: H = 127.05, p < 0.001).

**Figure 2.**
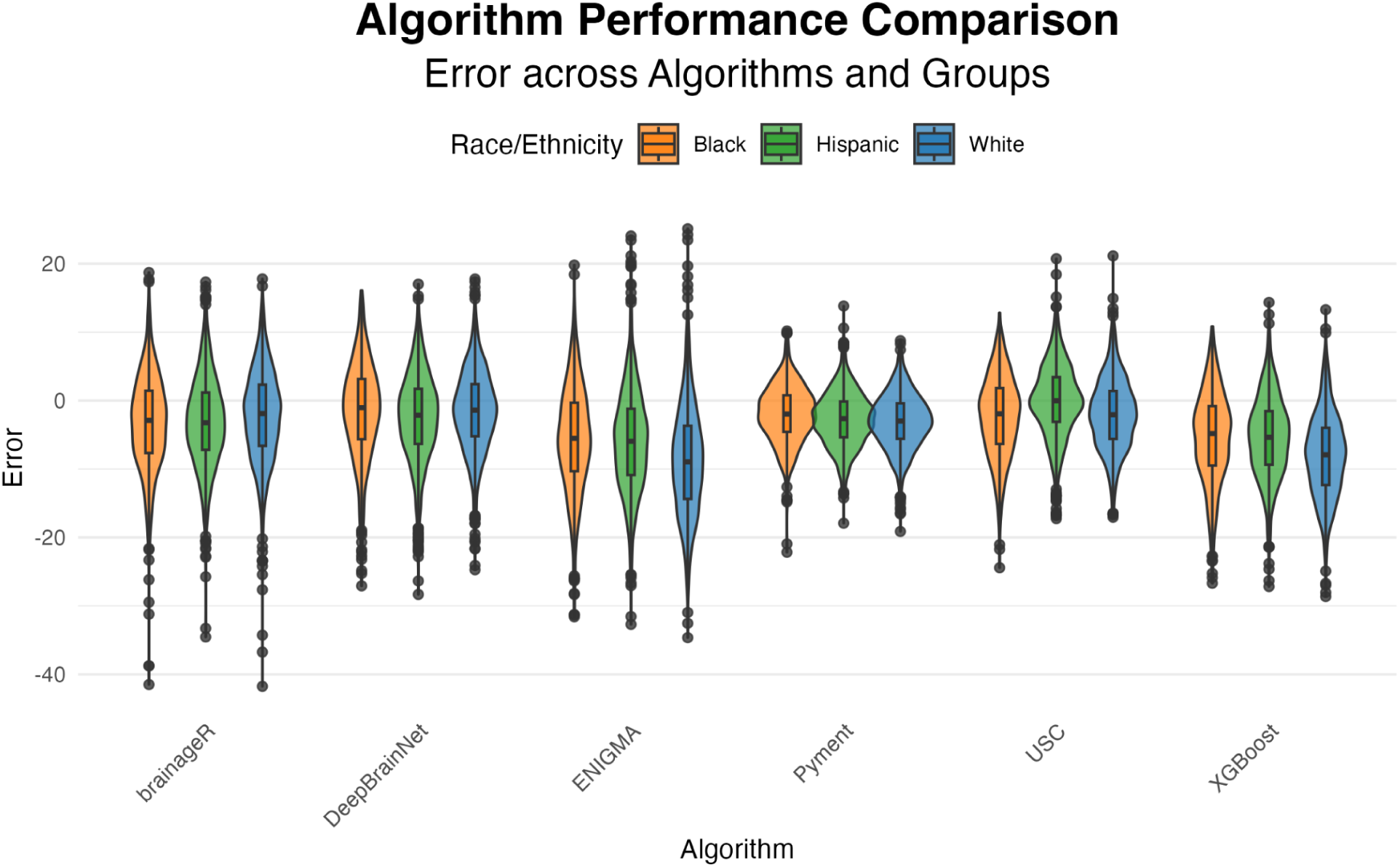
Error distributions for brainageR, DeepBrainNet, ENIGMA, USC, and XGBoost, depicted separately for Black participants (orange), Hispanic participants (green), and White participants (blue).

**Table 3.**
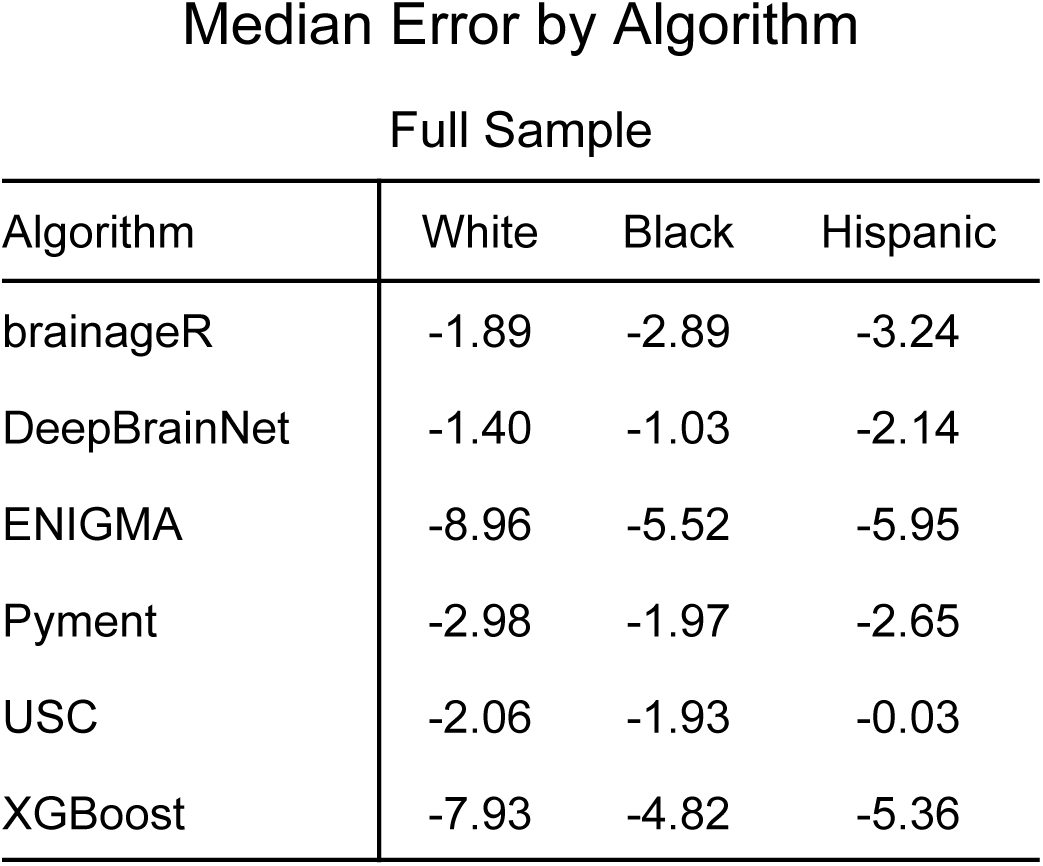
Median Error for each racial/ethnic group and algorithm in the full sample.

Post-hoc tests, however, revealed inconsistent–and in some instances, contrasting–patterns of intergroup differences across algorithms. Greater error in brain age prediction for African American compared to White American participants was found only for brainageR (z =-3.24, p-uncorrected=0.001, p-fdr= 0.002), whereas three algorithms had greater error for White American compared to African American participants; these were ENIGMA (z=8.76, p-uncorrected < 0.001, p-fdr < 0.001), Pyment (z=5.40, p-uncorrected < 0.001, p-fdr < 0.001), and XGBoost (z=9.20, p-uncorrected < 0.001, p-fdr < 0.001). In contrast, no differences in median error were found between African American and White American participants for DeepBrainNet (z=0.73) or USC (z=-0.31).

Comparing error across algorithms for White American and Hispanic American participants, significant differences between the two groups were found for DeepBrainNet; brainageR; ENIGMA; XGBoost; and USC. Specifically, White American median error was significantly greater than Hispanic American median error for ENIGMA (z =8.74, p-uncorrected < 0.001, p-fdr < 0.001), XGBoost (z =9.89, p-uncorrected < 0.001; p-fdr < 0.001), and USC (z=9.01, p-uncorrected < 0.001, p-fdr < 0.001), while Hispanic American median error was greater than White American for DeepBrainNet (z =-2.93, p-uncorrected= 0.003, p-fdr = 0.005) and brainageR (z=-3.55, p-uncorrected < 0.001, p-fdr < 0.001). Median error did not differ between White American and Hispanic American participants for Pyment only (z=1.55).

Median errors were found to differ significantly between African American and Hispanic American participants for three of six algorithms. Hispanic American error was greater than African American error for DeepBrainNet (z=3.27, p-uncorrected=0.001, p-fdr=0.002) and also for Pyment (z=4.04, p-uncorrected < 0.001, p-fdr < 0.001). African American median error was found to be significantly greater than Hispanic American for USC only (z=-8.14, p-uncorrected < 0.001, p-fdr < 0.001). There were no differences in median error between African American and Hispanic American participants for brainageR, ENIGMA, or XGBoost (z=-0.15-1.14).

### Analyses in Propensity Weighted Sample

Weighted correlations between brain age and chronological age for all algorithms ranged from 0.58-.89 for White Americans; 0.51-0.85 for African Americans; and 0.57-0.88 for Hispanic Americans. As also seen in the full sample, across algorithms, there was a consistent trend of weaker correlations between brain age and chronological age for African Americans compared to White or Hispanic Americans. In fact, after propensity score weighting, all significant differences in brain age-chronological age correlations were between African American participants and White or Hispanic American participants; there were no significant differences between correlation coefficients for White and Hispanic Americans for any of the six algorithms examined (z=-1.26-1.46).

Weighted brain age-chronological age correlations were found to differ significantly between White Americans and African Americans for brainageR (p-uncorrected < 0.001; p-fdr < 0.001; z=4.06, CI=0.048-0.142); Pyment (p-uncorrected=0.007; p-fdr=0.021; z=2.69, CI=0.008-0.054); XGBoost (p-uncorrected=0.012; p-fdr=0.028; z=2.51, CI=0.015-0.123); and USC (p-uncorrected < 0.001; p-fdr<0.001; z=4.26, CI=0.048-0.136). Significant differences in weighted correlation coefficients between White American and African American participants did not survive FDR correction for ENIGMA only (p-uncorrected=0.036; p-fdr=0.064; z=2.10, CI=0.005-0.142). No significant differences in brain age-chronological age correlations were detected between White Americans and African Americans for DeepBrainNet (z=1.28).

Comparing Hispanic and African American participants, significant differences were found for DeepBrainNet (p-uncorrected=0.018; p-fdr=0.036; z=2.37, CI=0.008-0.091); brainageR (p-uncorrected < 0.001; p-fdr=0.001; z=3.76, CI=0.031-0.148); ENIGMA (p-uncorrected=0.006; p-fdr=0.021; z=2.74, CI=0.026-0.162); XGBoost (p-uncorrected=0.009; p-fdr=0.024; z=2.60, CI=0.017-0.126); and USC (p-uncorrected=0.003; p-fdr=0.013; z=2.98, CI=0.022-0.113). There were no significant differences detected between Hispanic Americans and African Americans for Pyment (z=1.83).

Examining performance error, results found in the propensity weighted sample were similar to those reported in the full sample. For all algorithms except for XGBoost (H=2.00, p = 0.065), weighted Kruskal-Wallis tests indicated significant differences between the weighted median errors of the three racial/ethnic groups (all H’s = 2.00; brainageR: p < 0.001; DeepBrainNet: p < 0.001; ENIGMA: p = 0.016; Pyment: p = 0.015; USC: p < 0.001). All weighted median errors for all algorithms and groups are shown in Table 4.

**Table 4.**
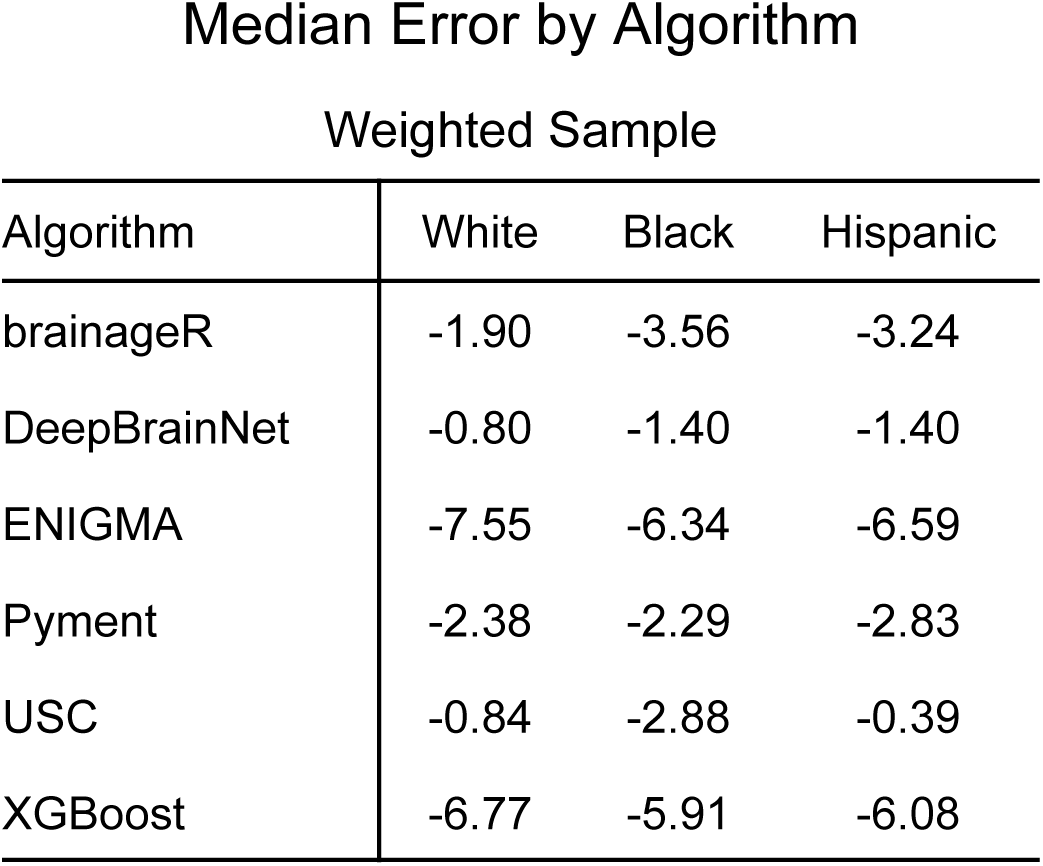
Weighted median error for each racial/ethnic group and algorithm in the propensity weighted sample.

Across algorithms, post-hoc tests again revealed highly variable patterns of cross-ethnoracial differences in weighted median error. Four algorithms–-DeepBrainNet, brainageR, ENIGMA, and USC–were all found to have weighted median errors that significantly differed between African American and White American participants. In contrast to the full sample, three algorithms had greater error for African American than for White American participants; specifically, African American error was greater than White American error for brainageR (p-uncorrected < 0.001; p-fdr <0.001; W=-4.52), USC (p-uncorrected < 0.001; p-fdr < 0.001; W=-6.98), and DeepBrainNet (p-uncorrected=0.038; p-fdr=0.06; W=-2.07), although the latter did not survive correction for multiple comparisons. African American error was lower than White American error for ENIGMA only (p-uncorrected=0.029; p-fdr= 0.049; W=2.18). No significant differences were found between African American and White American median errors for Pyment (W=0.48).

Between White Americans and Hispanic Americans, weighted median errors also significantly differed for four of six algorithms. Weighted median error was greater for Hispanic Americans than for White Americans for DeepBrainNet (p-uncorrected < 0.001; p-fdr < 0.001; W=-4.80); brainageR (p-uncorrected < 0.001; p-fdr < 0.001; W=-3.66); and Pyment (p-uncorrected=0.014; p-fdr=0.029; W=-2.45), and lower for Hispanic Americans than for White Americans for ENIGMA only (p-uncorrected=0.008; p-fdr=0.020; W=2.65). For USC post-hoc only, no significant difference was found between White American and Hispanic American weighted median errors (W=1.52).

Comparing Hispanic Americans and African Americans, significant differences were found for Pyment and USC only. Specifically, African American error was greater than Hispanic American error for USC (p-uncorrected < 0.001; p-fdr < 0.001; W=8.06), while Hispanic American error was greater than African American error for Pyment (p-uncorrected=0.016; p-fdr=0.029; W=-2.42). No differences between Hispanic American and African American weighted median errors were detected for DeepBrainNet, brainageR, or ENIGMA (z=-1.89-1.46). Altogether, and very similarly to the full sample, this pattern of results indicates that while most algorithms differentially predicted brain age based on race/ethnicity, the specific pattern of intergroup differences varied between them despite balancing age, sex, and Freesurfer Euler Number across groups.

## DISCUSSION

Here, we examined multiple indices of performance for the derivation of brain age in White-, African-, and Hispanic-American participants. Correlations between brain age and chronological age, as well as error between these two variables, were calculated for six commonly used brain age algorithms. To minimize the effect of confounding variables, we also completed similar analyses in a propensity score weighted sample. In the full sample, for most algorithms tested, the correlations between chronological age and brain age estimates were weaker for African Americans than for Hispanic Americans and/or White Americans. Median error differed by racial/ethnic group for all algorithms examined. In the propensity weighted sample, differences in correlations between brain age and chronological age persisted for the majority of algorithms after balancing age, sex, and a metric of scan quality between our racial and ethnic groups, while median error differed between racial/ethnic groups for all algorithms except for XGBoost. Our work begins to fill in important knowledge gaps, as brain age is becoming increasingly popular to probe acceleration in biological aging. However, the majority of brain age algorithms use data primarily from Non-Hispanic White subjects, although it is broadly understood that an algorithm’s training dataset may have implications for its clinical utility.

Connected to brain age, we believe that this is one of the first projects to systematically compare brain age predictions for multiple racial/ethnic groups. While past studies (Irajpour et al., 2025; Piçarra & Glocker, 2023) have examined racial/ethnic biases in brain age prediction, they have either primarily focused on algorithms infrequently used in applied research (e.g., ResNet-34) or they have tended to not directly test whether error levels differ between groups. Nonetheless, our findings regarding differences in correlations for African-American participants compared to other demographic groups agree with existing prior work (Li et al., 2022; Wisch et al., 2025). Past work has found that behavioral prediction was better for White Americans, compared to African-Americans, when functional MRI training datasets were composed of primarily White Americans (J. Li et al., 2022). Interestingly, even when models were trained on data only from African-Americans, behavioral prediction for African-Americans improved, but was still below levels seen for White Americans.

Other projects have sometimes reported older brain age in White-American versus African-American or Hispanic-American participants, finding that brain age was more sensitive to cognitive impairment for White-American participants compared to Hispanic- and African-American participants (Wisch et al., 2025). This is a slightly different question than we focused on here, as our group probed intra-individual variations (brain-v. chronological-age) between groups in six algorithms, as opposed to purely between-group differences in only two algorithms (DeepBrainNet; brainageR). Nonetheless, this prior work connects with some of our findings, such as larger error levels for African-American and Hispanic-American participants compared to White-American participants in the propensity weighted sample. If brain age is a biomarker for cognitive impairment, having greater errors in certain groups may translate into less sensitivity when trying to find pathology or abnormality in those same groups.

One critical piece to highlight is our use of error as a performance metric, in place of absolute error or squared error. Despite potentially causing some variability in results compared to past projects, this was motivated by several factors. First, neither absolute error nor squared error maintains the directionality of a discrepancy when judging performance of brain age predictions in relation to chronological age. Under v. over-estimations of brain age have distinct clinical implications, and as such, we feel that this is an important piece to consider in future work probing brain age. Second, many past projects simply report mean absolute or squared error, but rarely statistically test these metrics. This is a major limitation and important to highlight. The mean absolute or squared error of different groups could overlap with each other if individual error is sufficiently large and there is high variance for certain groups. Furthermore, the usage of absolute error or squared error in commonly used statistical models (e.g., ANOVAs; linear regression) is likely to violate different assumptions needed for these statistical tests. For example, linear models presume heteroscedasticity of residuals, but absolute error or squared error are not normal (Gaussian) in their distributional properties. Therefore, by using error as our primary metric, we were better able to capture important patterns in brain age prediction accuracy across racial and ethnic groups while completing rich and valid statistical modeling to formally test for potential algorithmic biases.

Thinking about our results, it is not clear why these differences might be emerging. One of the strongest potential explanations is related to the racial and ethnic composition of the training datasets. Many of the algorithms examined here were trained on similar, mostly racially and ethnically homogenous MRI cohorts, including: the UK Biobank (used in XGBoost, Pyment, and USC); the Autism Brain Imaging Data Exchange (used in brainageR, XGBoost, and Pyment); the Cambridge Centre for Ageing and Neuroscience (used in XGBoost, Pyment, and USC); the Human Connectome Project (used in XGBoost, Pyment, and USC); and the Pediatric Imaging, Neurocognition, and Genetics Study (used in XGBoost, Pyment, and DeepBrainNet). Not all of these studies report the full racial and ethnic makeup of their MRI samples, but the UK Biobank, for example, is approximately 94% White.

Alternatively, there could be other variables affecting brain age that differ between racial and ethnic groups. These could include factors such as neurocognitive functioning, socioeconomic status, and cumulative life stress, and could help explain why some brain age algorithms performed particularly poorly in Hispanic-American, but not White- and African-American samples. Greater variation in one or more of these variables could be linked to greater structural fMRI variability in African-American or Hispanic-American participants, causing algorithms to estimate brain age differently in these groups compared to White-Americans. Finally, the inputs to the different brain age algorithms could drive variations in brain age prediction error. Freesurfer is used by ENIGMA, Pyment, USC, and XGBoost to calculate brain age, and it remains an open question whether cross-racial/ethnic variation in brain age error could be related to templates used in Freesurfer that are based on a small number of participants (∼64). Interestingly, the actual racial and ethnic composition of the individuals whose scans were used to create Freesurfer templates remains unknown. We reached out to the developers of Freesurfer and staff from the Open Access Series of Imaging Studies, but neither entity was able to provide this information.

As with all studies, this project was not without its limitations. First, we attempted to minimize the potential for confounding by utilizing propensity score weighting to balance age, sex, and Euler Number across racial/ethnic groups, but there could have been additional confounding from demographic, neurocognitive, and other factors. Future work could consider including additional cognitive, developmental, or demographic variables that were not included in our project. Second, we were interested in applied work with brain age algorithms that were deployed across diverse clinical settings with different scanners. As MRI data was collected on either SKYRA or VIDA 3T Siemens scanners, there is a great deal of variability in scanner and acquisition protocols used here that could have increased noise in brain age estimation (Reynolds et al., 2023; Yamashita et al., 2019). This noise may be due to biological sources or engineering measurement bias, but harmonization often reduces variance by presuming that scanner differences are simple effects that can be cleanly separated from biological signals. Such approaches may still be in need of work and refinement (Gebre et al., 2023).

Third, we did not interrogate if correlations or errors varied as a function of age. The HABS-HD project recruited participants that were aged 50 or older, and scatterplots (i.e., Figure 1) suggest different levels of error for younger and older participants in the cohort. While this question was outside the scope of our project, we believe that it could be more richly interrogated in a future study. Fourth, we only tested 6 algorithms, and new algorithms will continue to emerge as this field progresses. For example, the DunedinPACNI estimates the longitudinal Pace of Aging from a single brain image (Whitman et al., 2025), but it was published after we had derived our measures of interest. Finally, this project was cross-sectional in nature, and therefore it will be important to examine if brain age prediction accuracy changes over time within individuals across different racial and ethnic groups. This could be a critical piece of knowledge, especially as we think about generalizability and the clinical utility of brain age. Despite these limitations, the results of this work illuminate gaps in the existing brain age literature while also generating many important questions to explore in future research. We were particularly interested to know if the level of bias present in different algorithms was related to the racial and ethnic composition of training datasets, but the majority of brain age algorithms examined were not accompanied by this information in different publications. To correct this issue, we would mandate that biological age algorithm developers detail the racial/ethnic composition of their training set in future publications.

In conclusion, this work contributes to the emerging field of brain age research by probing differences in performance metrics by race/ethnicity in six popular brain age algorithms. In exploring these differences, this work raises questions about biases that may be unwittingly perpetuated in brain age or other biological age algorithms; how these biases may be introduced into the development of an algorithm; and how they may be reduced or mitigated. Our results may intuitively lead to speculation that training algorithms on specific demographic groups should resolve this issue, yet nascent research suggests that algorithms trained on specific groups also show these biases, though to a lesser degree (J. Li et al., 2022). Therefore, evidence of these differences challenges us to explore more hidden ways in which these biases may be introduced, such in the sample recruitment, data collection, or MRI preprocessing stages. Ultimately, as these algorithms are already being utilized in clinical settings, small but significant differences in the estimates that these algorithms generate for different individuals can, on a larger scale, contribute to differences in clinical diagnosis, treatment, and care. The results of this work, therefore, are an important step towards furthering the goals of unbiased neuroscientific research as well as equity in neurocognitive healthcare.

## Acknowledgments

This work was supported by the National Institute of Mental Health under grant R21MH128793 to Dr. Hanson, as well as internal funding from the University of Pittsburgh and Pitt’s Learning Research & Development Center provided to Dr. Hanson. We thank Drs. Aaron Heller and Charles DeCarli for thoughtful discussion about brain age across different racial and ethnic groups.

## Ethical Information

The Health & Aging Brain Study obtained informed consent from participants in accordance with the University of North Texas Health Science Center Institutional Review Board. Our work (data analyses of de-identified data) was determined to not meet the definition of human subjects research, as defined by the U.S. Department of Health and Human Services and Food and Drug Administration regulations.

## Data Availability Statement

Data used in this study were obtained from the Health and Aging Brain Study - Health Disparities (https://apps.unthsc.edu/itr/our), and is publicly available to eligible researchers by request.

## Declaration of interests

The authors have declared that they have no competing or potential conflicts of interest.

## Notes

### Competing Interest Statement

The authors have declared no competing interest.

### Author Declarations

The HABS-HD study was approved by the University of North Texas Health Science Center Institutional Review Board. The Health & Aging Brain Study obtained informed consent from participants in accordance with the University of North Texas Health Science Center Institutional Review Board. Our work (data analyses of deidentified data) was determined to not meet the definition of human subjects research, as defined by the U.S. Department of Health and Human Services and Food and Drug Administration regulations.

## References

Bacas, E., Kahhalé, I., Raamana, P. R., Pablo, J. B., Anand, A. S., & Hanson, J. L. (2023). Probing multiple algorithms to calculate brain age: Examining reliability, relations with demographics, and predictive power. Human Brain Mapping, 44(9), 3481–3492.

Baecker, L., Garcia-Dias, R., Vieira, S., Scarpazza, C., & Mechelli, A. (2021). Machine learning for brain age prediction: Introduction to methods and clinical applications. EBioMedicine, 72.

Bashyam, V. M., Erus, G., Doshi, J., Habes, M., Nasrallah, I. M., Truelove-Hill, M., Srinivasan, D., Mamourian, L., Pomponio, R., Fan, Y., & others. (2020). MRI signatures of brain age and disease over the lifespan based on a deep brain network and 14 468 individuals worldwide. Brain, 143(7), 2312–2324.

Chen, R. J., Wang, J. J., Williamson, D. F., Chen, T. Y., Lipkova, J., Lu, M. Y., Sahai, S., & Mahmood, F. (2023). Algorithmic fairness in artificial intelligence for medicine and healthcare. Nature Biomedical Engineering, 7(6), 719–742.

Clark, A. L., Thomas, K. R., Ortega, N., Haley, A. P., Duarte, A., O’Bryant, S., & Team, for the H.-H. S. (2024). Empirically derived psychosocial-behavioral phenotypes in Black/African American and Hispanic/Latino older adults enrolled in HABS-HD: Associations with AD biomarkers and cognitive outcomes. Alzheimer’s & Dementia, 20(2), 1360–1373. 10.1002/alz.13544

Cole, J. H., Marioni, R. E., Harris, S. E., & Deary, I. J. (2019). Brain age and other bodily ‘ages’: Implications for neuropsychiatry. Molecular Psychiatry, 24(2), 266–281.

Cole, J. H., Ritchie, S. J., Bastin, M. E., Hernández, V., Muñoz Maniega, S., Royle, N., Corley, J., Pattie, A., Harris, S. E., Zhang, Q., & others. (2018). Brain age predicts mortality. Molecular Psychiatry, 23(5), 1385–1392.

Daneshjou, R., Vodrahalli, K., Novoa, R. A., Jenkins, M., Liang, W., Rotemberg, V., Ko, J., Swetter, S. M., Bailey, E. E., Gevaert, O., & others. (2022). Disparities in dermatology AI performance on a diverse, curated clinical image set. Science Advances, 8(31), eabq6147.

Dempsey, D. A., Deardorff, R., Saykin, A. J., & Risacher, S. L. (2023). BrainAGE methods: Influence of field strength, voxel size, race, and ethnicity. Alzheimer’s & Dementia, 19, e079921.

Dörfel, R. P., Arenas-Gomez, J. M., Fisher, P. M., Ganz, M., Knudsen, G. M., Svensson, J. E., & Plavén-Sigray, P. (2023). Prediction of brain age using structural magnetic resonance imaging: A comparison of accuracy and test–retest reliability of publicly available software packages. Human Brain Mapping, 44(17), 6139–6148.

Esteva, A., Kuprel, B., Novoa, R. A., Ko, J., Swetter, S. M., Blau, H. M., & Thrun, S. (2017). Dermatologist-level classification of skin cancer with deep neural networks. Nature, 542(7639), 115–118.

Fischl, B. (2012). FreeSurfer. Neuroimage, 62(2), 774–781.

Gebre, R. K., Senjem, M. L., Raghavan, S., Schwarz, C. G., Gunter, J. L., Hofrenning, E. I., Reid, R. I., Kantarci, K., Graff-Radford, J., Knopman, D. S., & others. (2023). Cross–scanner harmonization methods for structural MRI may need further work: A comparison study. NeuroImage, 269, 119912.

Gilmore, A. D., Buser, N. J., & Hanson, J. L. (2021). Variations in structural MRI quality significantly impact commonly used measures of brain anatomy. Brain Informatics, 8, 1–15.

Glasser, M. F., Coalson, T. S., Robinson, E. C., Hacker, C. D., Harwell, J., Yacoub, E., Ugurbil, K., Andersson, J., Beckmann, C. F., Jenkinson, M., & others. (2016). A multi-modal parcellation of human cerebral cortex. Nature, 536(7615), 171–178.

Hall, J. R., Petersen, M., Johnson, L., & O’Bryant, S. E. (2022). Characterizing Plasma Biomarkers of Alzheimer’s in a Diverse Community-Based Cohort: A Cross-Sectional Study of the HAB-HD Cohort. Frontiers in Neurology, 13, 871947.

Han, L. K., Dinga, R., Hahn, T., Ching, C. R., Eyler, L. T., Aftanas, L., Aghajani, M., Aleman, A., Baune, B. T., Berger, K., & others. (2021). Brain aging in major depressive disorder: Results from the ENIGMA major depressive disorder working group. Molecular Psychiatry, 26(9), 5124–5139.

Hanson, J. L., Adkins, D. J., Bacas, E., & Zhou, P. (2024). Examining the reliability of brain age algorithms under varying degrees of participant motion. Brain Informatics, 11(1), 9.

Hayashi, S., Caron, B. A., Heinsfeld, A. S., Vinci-Booher, S., McPherson, B., Bullock, D. N., Bertò, G., Niso, G., Hanekamp, S., Levitas, D., & others. (2024). Brainlife. Io: A decentralized and open-source cloud platform to support neuroscience research. Nature Methods, 21(5), 809–813.

Irajpour, M., Barekatain, M., Karami, M., Alavijeh, S. K., Barekatain, M., & Irajpour, M. (2025). Advanced Brain Age Prediction Using Multi-Head Self-Attention: A Comparative Analysis of Western and Middle Eastern MRI Datasets. Research Square, rs-3.

Kaufmann, T., van der Meer, D., Doan, N. T., Schwarz, E., Lund, M. J., Agartz, I., Alnæs, D., Barch, D. M., Baur-Streubel, R., Bertolino, A., & others. (2019). Common brain disorders are associated with heritable patterns of apparent aging of the brain. Nature Neuroscience, 22(10), 1617–1623.

Kleinberg, G., Diaz, M. J., Batchu, S., & Lucke-Wold, B. (2022). Racial underrepresentation in dermatological datasets leads to biased machine learning models and inequitable healthcare. Journal of Biomed Research, 3(1), 42.

Leonardsen, E. H., Peng, H., Kaufmann, T., Agartz, I., Andreassen, O. A., Celius, E. G., Espeseth, T., Harbo, H. F., Høgestøl, E. A., De Lange, A.-M., & others. (2022). Deep neural networks learn general and clinically relevant representations of the ageing brain. NeuroImage, 256, 119210.

Li, J., Bzdok, D., Chen, J., Tam, A., Ooi, L. Q. R., Holmes, A. J., Ge, T., Patil, K. R., Jabbi, M., Eickhoff, S. B., & others. (2022). Cross-ethnicity/race generalization failure of behavioral prediction from resting-state functional connectivity. Science Advances, 8(11), eabj1812.

Li, Z., Zhang, Z., Ren, Y., Wang, Y., Fang, J., Yue, H., Ma, S., & Guan, F. (2021). Aging and age-related diseases: From mechanisms to therapeutic strategies. Biogerontology, 22(2), 165–187.

Mather, K. A., Jorm, A. F., Parslow, R. A., & Christensen, H. (2011). Is telomere length a biomarker of aging? A review. Journals of Gerontology Series A: Biomedical Sciences and Medical Sciences, 66(2), 202–213.

O’Bryant, S. E., Johnson, L. A., Barber, R. C., Braskie, M. N., Christian, B., Hall, J. R., Hazra, N., King, K., Kothapalli, D., Large, S., Mason, D., Matsiyevskiy, E., McColl, R., Nandy, R., Palmer, R., Petersen, M., Philips, N., Rissman, R. A., Shi, Y., … Team, for the H. S. (2021). The Health & Aging Brain among Latino Elders (HABLE) study methods and participant characteristics. Alzheimer’s & Dementia: Diagnosis, Assessment & Disease Monitoring, 13(1), e12202. 10.1002/dad2.12202

Pal, S., & Tyler, J. K. (2016). Epigenetics and aging. Science Advances, 2(7), e1600584.

Philibert, R., Beach, S. R. H., Lei, M.-K., Gibbons, F. X., Gerrard, M., Simons, R. L., & Dogan, M. V. (2020). Array-Based Epigenetic Aging Indices May Be Racially Biased. Genes, 11(6), 685. 10.3390/genes11060685

Piçarra, C., & Glocker, B. (2023). Analysing race and sex bias in brain age prediction. Workshop on Clinical Image-Based Procedures, 194–204.

Reynolds, M., Chaudhary, T., Torbati, M. E., Tudorascu, D. L., Batmanghelich, K., Initiative, A. D. N., & others. (2023). Combat harmonization: Empirical bayes versus fully bayes approaches. NeuroImage: Clinical, 39, 103472.

Rosen, A. F., Roalf, D. R., Ruparel, K., Blake, J., Seelaus, K., Villa, L. P., Ciric, R., Cook, P. A., Davatzikos, C., Elliott, M. A., & others. (2018). Quantitative assessment of structural image quality. Neuroimage, 169, 407–418.

Turney, I. C., Lao, P. J., Rentería, M. A., Igwe, K. C., Berroa, J., Rivera, A., Benavides, A., Morales, C. D., Rizvi, B., Schupf, N., Mayeux, R., Manly, J. J., & Brickman, A. M. (2023). Brain Aging Among Racially and Ethnically Diverse Middle-Aged and Older Adults. JAMA Neurology, 80(1), 73–81. 10.1001/jamaneurol.2022.3919

Tustison, N. J., Cook, P. A., Holbrook, A. J., Johnson, H. J., Muschelli, J., Devenyi, G. A., Duda, J. T., Das, S. R., Cullen, N. C., Gillen, D. L., & others. (2021). The ANTsX ecosystem for quantitative biological and medical imaging. Scientific Reports, 11(1), 9068.

Whitman, E. T., Elliott, M. L., Knodt, A. R., Abraham, W. C., Anderson, T. J., Cutfield, N. J., Hogan, S., Ireland, D., Melzer, T. R., Ramrakha, S., & others. (2025). DunedinPACNI estimates the longitudinal Pace of Aging from a single brain image to track health and disease. Nature Aging, 1–18.

Wisch, J. K., Petersen, K., Millar, P. R., Abdelmoity, O., Babulal, G. M., Meeker, K. L., Braskie, M. N., Yaffe, K., Toga, A. W., O’Bryant, S., Ances, B. M., & the HABS-HD Study Team. (2025). Cross-Sectional Comparison of Structural MRI Markers of Impairment in a Diverse Cohort of Older Adults. Human Brain Mapping, 46(2), e70133. 10.1002/hbm.70133

Yamashita, A., Yahata, N., Itahashi, T., Lisi, G., Yamada, T., Ichikawa, N., Takamura, M., Yoshihara, Y., Kunimatsu, A., Okada, N., & others. (2019). Harmonization of resting-state functional MRI data across multiple imaging sites via the separation of site differences into sampling bias and measurement bias. PLoS Biology, 17(4), e3000042.

Yin, C., Imms, P., Cheng, M., Amgalan, A., Chowdhury, N. F., Massett, R. J., Chaudhari, N. N., Chen, X., Thompson, P. M., Bogdan, P., Irimia, A., the Alzheimer’s Disease Neuroimaging Initiative, Weiner, M. W., Aisen, P., Petersen, R., Weiner, M. W., Aisen, P., Petersen, R., Jack, C. R., … Simpson, D. M. (2023). Anatomically interpretable deep learning of brain age captures domain-specific cognitive impairment. Proceedings of the National Academy of Sciences, 120(2), e2214634120. 10.1073/pnas.2214634120

